# Comparing 5-year survival between surgeons within pathological stage after gastric cancer resection: a single-center cohort study (the SAH framework)

**DOI:** 10.64898/2026.06.02.26354716

**Authors:** Birendra Kumar Sah, Danping Gu, Chenjie Dong, Jian Li, Minmin Zhang, Rui Jin, Xiaoyang Li, Erzhen Chen

## Abstract

**Background:** Long-term survival and major postoperative morbidity matter most to patients after cancer surgery, yet surgeon-level outcomes are rarely audited within pathological stage. We evaluated the Surgical Assessment in Healthcare framework, a stage-specific dual-outcome audit, at one center.

**Methods:** We studied 690 consecutive curative gastrectomies for Stage I to III gastric adenocarcinoma performed in 2019 by eight high-volume gastrointestinal oncology consultants (complete 5-year follow-up). The primary cohort included 570 surgery-first patients; 120 neoadjuvant patients entered sensitivity analysis. Co-primary outcomes were 5-year overall survival and major postoperative morbidity. Expected outcomes came from stage-stratified models fitted to all other surgeons’ patients (adjusted for age, sex, tumor location, size, and grade), with exact 95% confidence intervals and two-sided Holm adjustment across 23 exploratory tests.

**Results:** Among 570 patients, 118 (20.7%) died within 5 years and 37 (6.5%) experienced major morbidity. Overall surgeon observed-to-expected mortality ratios ranged from 0.77 to 1.62; neither overall mortality nor major morbidity differed after adjustment. One group differed: surgeon S6 in Stage II (9 observed versus 2.3 expected deaths among 20 patients; ratio 3.96, exact 95% confidence interval 2.11 to 5.90; exploratory Holm-adjusted P = 0.004), persisting across sensitivity analyses. Stage III showed no variation. Detecting doubled mortality with 80% power requires about 11 expected deaths per prespecified group (21 Holm-adjusted).

**Conclusion:** Stage-specific surgeon auditing of 5-year survival and major morbidity is feasible from routine data, but a single center reliably detects only large differences; routine use requires multi-year or multi-center pooling.

## Introduction

Gastric cancer management varies widely between countries and centers. Guidelines differ on the role and timing of neoadjuvant chemotherapy, the indications for resection and the extent of lymphadenectomy [1–3]. The operating surgeon stands at every decisive point: judging operability, deciding whether to operate first or refer for neoadjuvant chemotherapy [4,5], choosing the extent of resection and lymphadenectomy and, with the final pathology in hand, directing adjuvant treatment and follow-up. Adherence to operative standards varies even between specialized centers [6], and differences between surgeons may persist into long-term survival [7].

Quality measurement in gastric cancer surgery has concentrated on hospital volume and short-term outcomes [8–10]. Surgeon-level comparison is established in cardiac surgery [11] and in programs such as NSQIP [12,13], but long-term survival after cancer surgery is rarely compared between individual surgeons, because pathological stage dominates prognosis and can easily overshadow differences in surgical care.

Three further problems make such comparisons fragile. First, when expected deaths are estimated from the surgeons being compared, the ratios balance to one in total, so one surgeon’s excess is necessarily another’s deficit [14–16]. Second, stage after neoadjuvant chemotherapy (ypTNM) is not comparable with pathological stage after surgery alone. Third, with few deaths per surgeon, ratios are dominated by chance, and many comparisons produce chance findings [17,18]. We developed the Surgical Assessment in Healthcare (SAH) framework to avoid internal benchmarking bias and account for small-sample uncertainty, and applied it to a consecutive single-center cohort evaluating both 5-year overall survival and major postoperative morbidity, to establish what such a comparison can detect within a single center and what routine use would require.

## Materials and Methods

### Study design, ethics and reporting

This retrospective cohort study was conducted in the Department of General Surgery, Gastrointestinal Surgery Unit, Ruijin Hospital, Shanghai Jiao Tong University School of Medicine, Shanghai, China. It was approved by the Ruijin Hospital Ethics Committee (approval no. 2025-594; 18 September 2025), which waived individual consent for the use of de-identified data, and was registered at ClinicalTrials.gov (NCT07180966) after the cohort had been treated (retrospective registration of an observational study). The eight surgeons are coded S1 to S8. None of the authors are among the eight operating surgeons. Individual informed consent was waived for this retrospective institutional quality audit. The study followed the Declaration of Helsinki [19] and is reported according to STROBE [20].

### Patients

We screened consecutive patients who underwent gastrectomy for gastric adenocarcinoma between 1 January and 31 December 2019 under the care of eight high-volume consultant surgeons dedicated to gastrointestinal malignancy (n = 780). Rather than including every surgeon across the institution, this cohort intentionally focused on experienced, high-volume gastrointestinal oncology consultants, each with more than 15 years of gastric cancer experience. Eligible patients were aged 18 years or older and underwent elective curative-intent resection. We excluded 34 patients with non-adenocarcinoma histology or other ineligibility, 50 with Stage IV (pM1) disease, 4 with a pathological complete response after neoadjuvant chemotherapy and 2 records from a single patient entered twice who was found on re-audit to have peritoneal (M1) metastasis, leaving 690 patients with Stage I–III disease (AJCC 8th edition) [21] (Supplementary Figure 1). A pathologist blinded to the operating surgeon reviewed all discrepancies between recorded node counts and N category before analysis; two patients, both receiving neoadjuvant chemotherapy, were reclassified from ypStage III to ypStage II.

The primary cohort comprised the 570 patients treated with surgery first, whose stage is pathological (pTNM). The 120 patients receiving neoadjuvant chemotherapy, whose stage is ypTNM, were included only in a sensitivity analysis (Supplementary Table 3). Standard lymphadenectomy was D2 according to contemporaneous Japanese guidelines [22]. Adjuvant chemotherapy was guided by medical oncologists according to standard guidelines, typically CAPOX or SOX [22,23].

### Outcomes

The co-primary outcomes of the SAH framework were death from any cause within 5 years (60 months) of surgery and major postoperative morbidity, defined as Clavien–Dindo grade IIIa or higher [24]. Survival status was ascertained to 28 July 2025; the shortest follow-up among survivors was 68.1 months, so 5-year vital status was known for every patient. Complete vital status ascertainment for patients lost to routine clinic follow-up was achieved in coordination with the Shanghai Hospital Quality Control Center through cross-matching with national population and mortality registries.

### Adjustment variables

All comparisons were made within pTNM stage (I, II and III). Within stage, expected risk was adjusted for characteristics present before treatment and recorded for every patient: age (restricted cubic spline with knots at the 10th, 50th and 90th percentiles (47, 63 and 75 years)), sex, tumor location (proximal third or whole stomach versus middle or distal), log-transformed tumor size (11 missing values [1.6%] imputed to the median of 30 mm) and histological grade (differentiated [reference], undifferentiated, or not assessable [Gx]). Decisions led by the surgeon, including extent of gastrectomy, lymphadenectomy and adjuvant chemotherapy, were not adjusted for, because they may be part of the surgeon’s effect on survival; instead, they are reported separately as care-process measures (Table 3). ASA grade, comorbidity, performance status and clinical stage were not available.

### The SAH framework

The core concept of the SAH framework is a concrete, stage-specific surgical quality audit anchored on the two outcomes that matter most to patients after curative cancer surgery: 5-year overall survival and major postoperative morbidity, which directly affects postoperative recovery and quality of life. Both co-primary outcomes are evaluated for each surgeon as observed-to-expected (O/E) ratios benchmarked against all other surgeons rather than combined into a single arbitrary composite score. For each surgeon, a Cox model stratified by stage (separate baseline hazards for Stages I, II and III) was fitted to the other seven surgeons’ patients and used to predict each of that surgeon’s patients’ probability of death within 5 years. Expected deaths (E) are the sum of these probabilities over a surgeon’s patients, or over the patients in one surgeon × stage cell, and observed deaths (O) are the deaths that occurred. Because a surgeon’s own patients never contribute to his or her expected deaths, O and E are independent and the ratios need not balance across surgeons. The O/E ratio is a ratio of 5-year risks. Its two-sided 95% CI and exact two-sided P value were calculated from the distribution of a sum of independent binary outcomes (Poisson-binomial), with no large-sample approximation (Supplementary File 1); the steps are summarized in Supplementary Table 6.

Cells with at least 10 patients in Stages II and III were evaluated (15 cells); Stage I cells (12 deaths among 241 patients) are reported descriptively. Across these 23 comparisons (8 surgeons overall and 15 surgeon-by-stage cells), a surgeon or cell was considered to differ from expected when its Holm-adjusted two-sided P value [25] was below 0.05.

### Robustness checks

Any difference was examined in six ways: (i) on the hazard scale, in which each patient’s expected count is the cumulative hazard over his or her own follow-up; (ii) in all 690 patients, with neoadjuvant chemotherapy as an additional adjustment variable; (iii) with the model stratified by substage (IIA, IIB, IIIA, IIIB, IIIC); (iv) restricted to patients with at least 16 retrieved lymph nodes to test sensitivity to understaging; (v) via 500-replicate stratified bootstrap resampling propagating uncertainty in both the peer-surgeon reference model and the target cell; and (vi) with empirical Bayes shrinkage, which pulls each ratio toward 1 in proportion to its imprecision and estimates how much variation between surgeons exceeds chance [17,18]. Shrinkage used a Poisson–gamma model fitted separately to surgeons, Stage II cells and Stage III cells.

### Secondary analyses

Major morbidity was compared in the same way, with expected events from a logistic model containing stage and the adjustment variables. For each surgeon in the primary surgery-first cohort we tabulated total gastrectomy, laparoscopic approach, node yield (median and proportion with at least 16 nodes [26]), R0 resection, major morbidity, institutional follow-up at the study hospital and recorded adjuvant chemotherapy in Stage II–III (both overall and among patients followed at the study hospital), alongside overall caseload and neoadjuvant rate across all 690 patients. To quantify statistical power in a single center, we calculated the expected deaths a group requires for 80% power to detect true O/E ratios of 3.0, 2.0 and 1.5 at both an unadjusted two-sided 5% level (single prespecified cell) and the step-1 two-sided Holm threshold across 23 comparisons (0.05/23).

### Prespecification and software

Because this was a retrospective observational audit without a prespecified confirmatory protocol, all hypothesis tests and Holm-adjusted P values should be interpreted as exploratory. Analyses were performed in Python 3.13 with NumPy, SciPy and pandas; statistical specifications are provided in Supplementary File 1, and full Python analysis scripts are available from the corresponding author upon reasonable request. All primary and supplementary results were independently cross-checked directly against the de-identified SPSS dataset using lifelines and statsmodels. Continuous data are given as median (IQR) and categorical data as n (%).

## Results

### Patients

Of 780 patients screened, 690 were included, of whom 570 (82.6%) were treated with surgery first (Table 1). In the surgery-first primary cohort (n = 570), 241 patients (42.3%) had Stage I, 136 (23.9%) Stage II and 193 (33.9%) Stage III disease; median age was 63 years (IQR 56–70). Within 5 years, 118 patients (20.7%) died: 12 in Stage I, 22 in Stage II and 84 in Stage III. The remaining 120 patients (17.4%) received neoadjuvant chemotherapy, of whom 63 (52.5%) died within 5 years (Table 1; Supplementary Table 3).

**Table 1.**
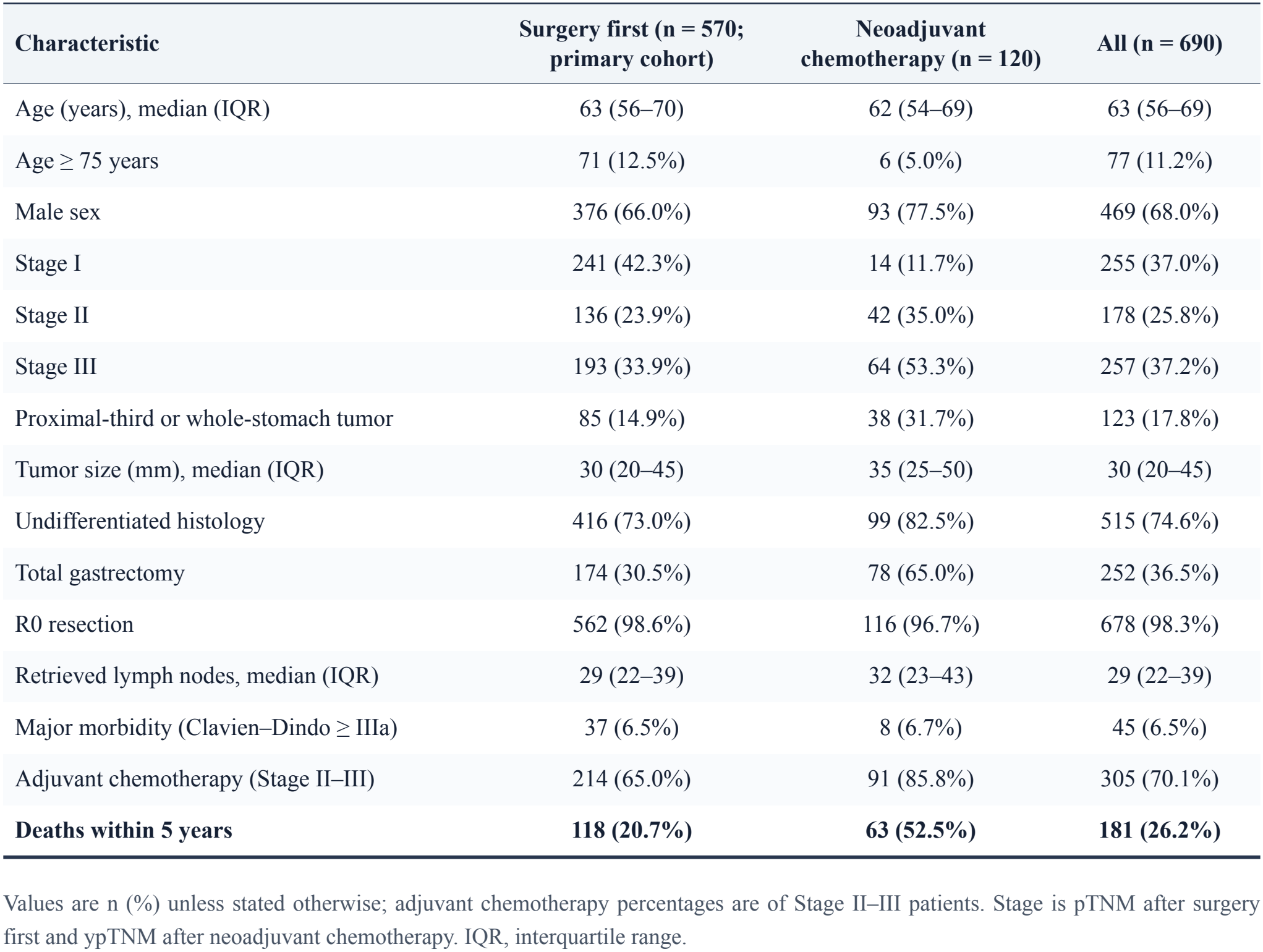
Patient characteristics and outcomes.

### Expected-risk model

In the surgery-first primary cohort, age was the only strong predictor within stage, and its effect was not linear (Supplementary Table 1): relative to the cohort median of 63 years, the modeled hazard at 50 years was similar (HR 0.95, 95% CI 0.70–1.29), whereas at 75 years (90th percentile) it was 2.48 times higher (1.87–3.29). The adjustment variables discriminated modestly within stage (Harrell’s C 0.75, 0.68 and 0.62 in Stages I, II and III). Expected deaths agreed with observed deaths in each stage (Stage I 12 observed vs 12.1 expected; Stage II 22 vs 21.9; Stage III 84 vs 83.6).

### Comparison between surgeons

In the surgery-first primary cohort (n = 570), surgeon-level O/E ratios ranged from 0.77 to 1.62 (Table 2). The highest was S6, with 22 deaths observed against 13.6 expected among 64 patients (O/E 1.62, 95% CI 1.14–2.11), but no surgeon differed after Holm adjustment for multiple testing (S6 Holm-adjusted P = 0.20). After empirical Bayes shrinkage, every surgeon-level ratio lay between 0.99 and 1.02, indicating that almost all overall variation between surgeons was compatible with chance.

**Table 2.** The SAH framework: observed and expected deaths within 5 years and O/E ratios by surgeon and stage (surgery-first cohort, n = 570)

| **Surgeon (total n)** | **Surgery-first (Stage I / II / III)** | **All stages** | **Stage II** | **Stage III** |
|---|---|---|---|---|
| S1 (139) | 110 (49 / 26 / 35) | 18 / 23.4<br>0.77 (0.50–1.08) | 3 / 5.0<br>0.60 (0.13–1.44) | 11 / 16.5<br>0.67 (0.37–1.01) |
| S2 (132) | 102 (41 / 30 / 31) | 17 / 20.8<br>0.82 (0.51–1.19) | 3 / 5.1<br>0.59 (0.13–1.53) | 12 / 13.4<br>0.89 (0.51–1.31) |
| S3 (109) | 94 (41 / 17 / 36) | 22 / 21.3<br>1.03 (0.71–1.38) | 2 / 3.1<br>0.65 (0.08–1.88) | 19 / 16.4<br>1.16 (0.80–1.51) |
| S4 (79) | 57 (25 / 13 / 19) | 10 / 12.2<br>0.82 (0.43–1.31) | 2 / 2.0<br>1.01 (0.13–2.88) | 8 / 8.7<br>0.92 (0.45–1.43) |
| S5 (66) | 54 (28 / 12 / 14) | 9 / 9.0<br>1.01 (0.51–1.61) | 1 / 1.7<br>0.59 (0.02–2.57) | 6 / 6.2<br>0.97 (0.41–1.58) |
| S6 (66) | 64 (20 / 20 / 24) | 22 / 13.6<br>1.62 (1.14–2.11) | **9 / 2.3<br>3.96 (2.11–5.90) \*** | 11 / 9.7<br>1.14 (0.65–1.64) |
| S7 (56) | 49 (20 / 11 / 18) | 10 / 8.6<br>1.17 (0.61–1.86) | 1 / 1.5<br>0.65 (0.02–2.87) | 8 / 6.1<br>1.31 (0.64–2.03) |
| S8 (43) | 40 (17 / 7 / 16) | 10 / 8.8<br>1.14 (0.62–1.74) | 1 / 1.3<br>0.79 (0.02–3.07)<sup>a</sup> | 9 / 6.7<br>1.35 (0.74–1.90) |

Surgeons are coded S1 to S8 (total caseload across all 690 patients is shown in parentheses). Each entry shows observed / expected deaths, followed by O/E (exact two-sided 95% CI). Expected deaths come from a stage-stratified Cox model fitted to the other seven surgeons' patients. Values in bold indicate a statistically significant excess (\* differs from expected after two-sided Holm adjustment across 23 exploratory tests, P = 0.004). <sup>a</sup> Fewer than 10 patients; descriptive only. Stage I cells are shown in Supplementary Table 2.

Of the 15 evaluable surgeon × stage cells, one differed from expected: S6 in Stage II, with 9 deaths observed against 2.27 expected among 20 patients (O/E 3.96, 95% CI 2.11–5.90; Holm-adjusted P = 0.004) (Table 2; Fig. 1). In Stage III, cell ratios ranged from 0.67 to 1.35, none differed, and empirical Bayes shrinkage brought every Stage III cell to 1.00, indicating no detectable variation between surgeons (Fig. 2); an unadjusted log-rank test across all eight surgeons gave P = 0.71 in Stage III and P = 0.012 in Stage II.

**Figure 1.**
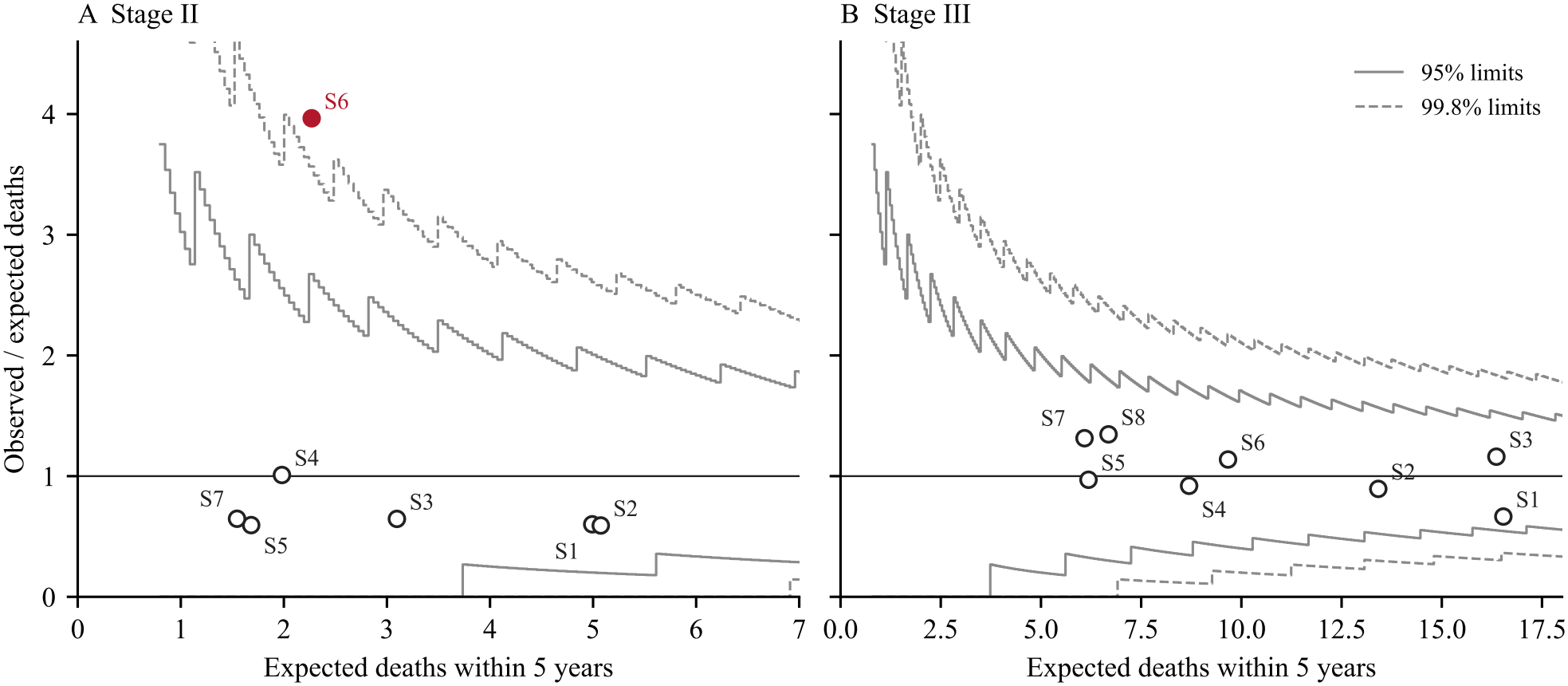
Funnel plot of observed-to-expected (O/E) 5-year mortality ratios plotted against expected deaths for each evaluable surgeon-by-stage group in (A) Stage II and (B) Stage III (surgery-first cohort, n = 570). Expected deaths come from a stage-stratified Cox model fitted to the other seven surgeons’ patients. Solid and dashed lines indicate exact Poisson 95% and 99.8% control limits. The crimson point marks S6 in Stage II (9 observed vs 2.27 expected deaths; O/E 3.96, Holm-adjusted P = 0.004), the only group that differed from expected after Holm adjustment across 23 tests.

**Figure 2.**
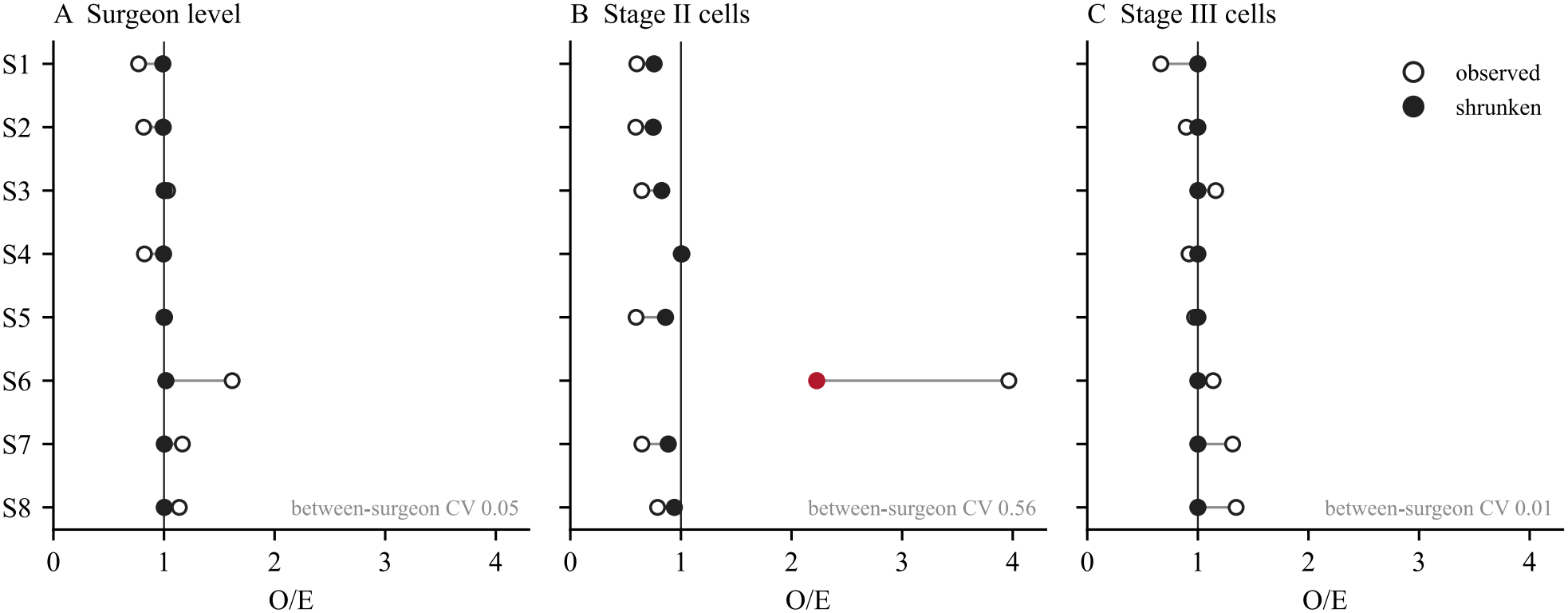
Observed and empirical Bayes shrunken observed-to-expected (O/E) 5-year mortality ratios across (A) surgeons overall, (B) Stage II, and (C) Stage III (surgery-first cohort, n = 570). Open circles show observed O/E ratios and filled circles show Poisson–gamma empirical Bayes shrunken estimates; the crimson filled circle marks S6 in Stage II (shrunken O/E 2.23; posterior probability 0.99 that the true ratio exceeds 1). Estimated between-surgeon coefficients of variation (CV) are shown in each panel. The vertical reference line indicates O/E = 1.0.

### Robustness

The S6 × Stage II result was consistent in every check (Supplementary Table 5): O/E 4.12 (1.89–7.83) on the hazard scale, 3.29 (1.81–4.82) in all 690 patients, 4.14 (2.30–6.03) with substage stratification and 3.78 (1.92–5.67; 8 observed vs 2.11 expected in n = 17) when restricted to patients with at least 16 retrieved nodes. Propagating reference-model uncertainty via 500-replicate bootstrap widened the 95% CI to 1.69– 8.67 (expected deaths 95% CI 1.13–3.86), remaining above 1. After empirical Bayes shrinkage, the ratio was 2.23, with a posterior probability of 0.99 that the true ratio exceeded 1. S6’s Stage II patients did not have a higher proportion of Stage IIB disease (11 of 20 [55.0%] vs 68 of 116 [58.6%] for the other surgeons).

The second-highest cell, S8 in Stage III (O/E 1.35, 95% CI 0.74–1.90), had a 95% CI above 1 only on the hazard scale (O/E 2.29, 95% CI 1.05–4.35). This reflects earlier timing of death rather than a higher 5-year death count: S8’s Stage III deaths occurred earlier (median 15.1 vs 28.6 months for the other surgeons), and a patient who dies early accrues less expected cumulative hazard, lowering expected deaths on the hazard scale (3.9 vs 6.7 on the 5-year risk scale) while observed deaths remain unchanged.

### Major morbidity and care-process measures

Major morbidity occurred in 37 patients (6.5%) against 36.4 expected; no surgeon or cell differed, but with fewer than four expected events per cell, this analysis lacked adequate statistical power to detect even a twofold increase in complication risk (Supplementary Table 4).

In the surgery-first cohort (Table 3), S6 had the lowest proportion of patients with at least 16 retrieved lymph nodes (76.6% vs 87.0–98.2%; median 23.5 nodes) and the lowest neoadjuvant chemotherapy rate across all 690 patients (3.0% vs 7.0–27.8%). S6 had the lowest overall recorded adjuvant chemotherapy rate in Stage II–III (47.7% vs 52.5–88.7%), and this coincided with the lowest institutional follow-up rate at the study hospital (50.0% vs 59.4–91.8%); among Stage II–III patients followed at the study hospital, S6’s adjuvant chemotherapy rate was 86.4% (9 of 10 [90.0%] in Stage IIB vs 64.7% for the other surgeons), meaning that incomplete capture of outside-hospital chemotherapy cannot be separated from lower treatment delivery outside the study hospital. R0 resection (98.4%) and major morbidity (7.8%) were similar to those of the other surgeons. S6 had the second-highest proportion of patients aged 75 years or older (17.2%; S8 17.5%; other surgeons 5.6–14.7%).

**Table 3.**
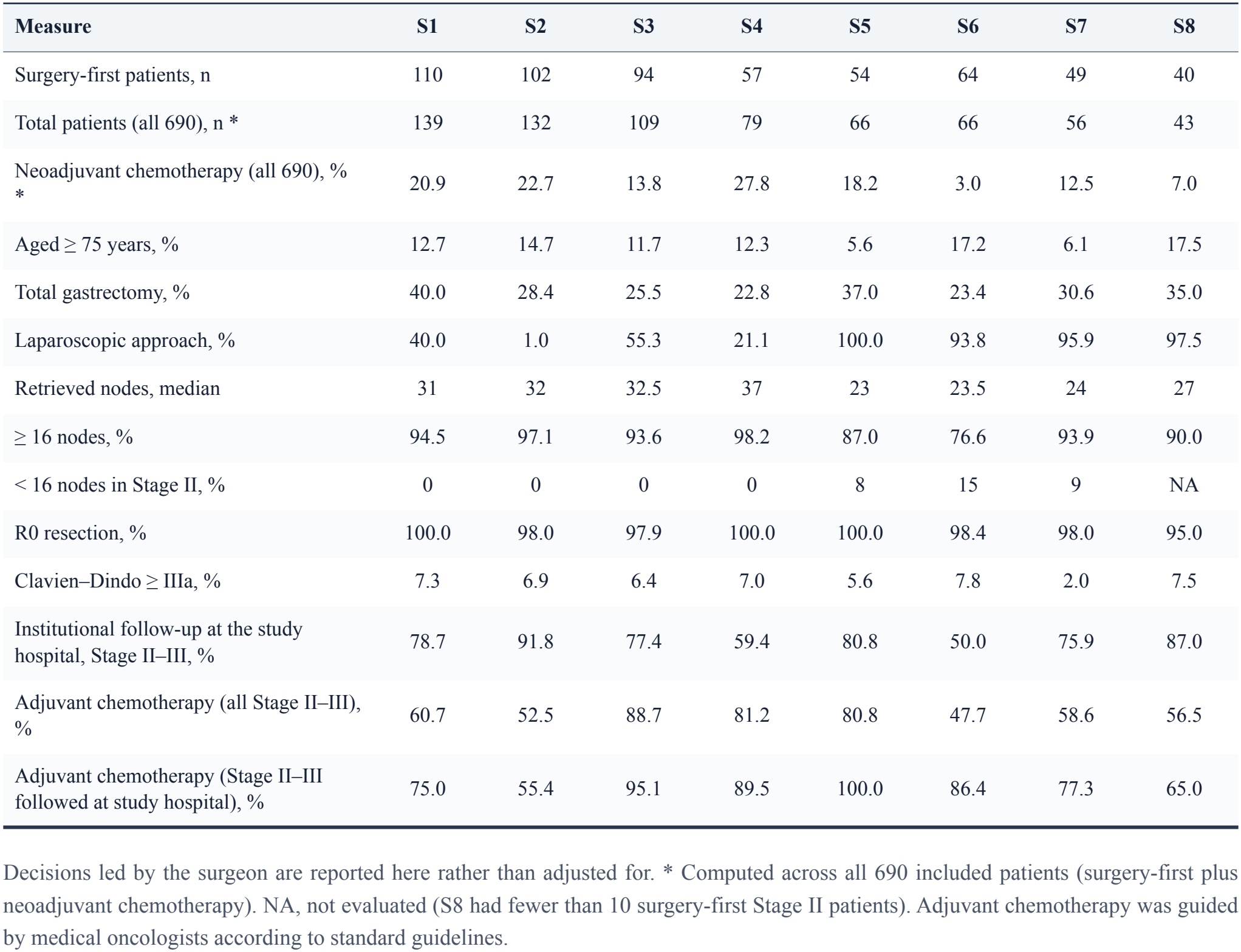
Process measures and case mix by surgeon (surgery-first primary cohort, n = 570)

| **Measure** | **S1** | **S2** | **S3** | **S4** | **S5** | **S6** | **S7** | **S8** |
|---|---|---|---|---|---|---|---|---|
| Surgery-first patients, n | 110 | 102 | 94 | 57 | 54 | 64 | 49 | 40 |
| Total patients (all 690), n * | 139 | 132 | 109 | 79 | 66 | 66 | 56 | 43 |
| Neoadjuvant chemotherapy (all 690), % * | 20.9 | 22.7 | 13.8 | 27.8 | 18.2 | 3.0 | 12.5 | 7.0 |
| Aged ≥ 75 years, % | 12.7 | 14.7 | 11.7 | 12.3 | 5.6 | 17.2 | 6.1 | 17.5 |
| Total gastrectomy, % | 40.0 | 28.4 | 25.5 | 22.8 | 37.0 | 23.4 | 30.6 | 35.0 |
| Laparoscopic approach, % | 40.0 | 1.0 | 55.3 | 21.1 | 100.0 | 93.8 | 95.9 | 97.5 |
| Retrieved nodes, median | 31 | 32 | 32.5 | 37 | 23 | 23.5 | 24 | 27 |
| ≥ 16 nodes, % | 94.5 | 97.1 | 93.6 | 98.2 | 87.0 | 76.6 | 93.9 | 90.0 |
| < 16 nodes in Stage II, % | 0 | 0 | 0 | 0 | 8 | 15 | 9 | NA |
| R0 resection, % | 100.0 | 98.0 | 97.9 | 100.0 | 100.0 | 98.4 | 98.0 | 95.0 |
| Clavien–Dindo ≥ IIIa, % | 7.3 | 6.9 | 6.4 | 7.0 | 5.6 | 7.8 | 2.0 | 7.5 |
| Institutional follow-up at the study hospital, Stage II–III, % | 78.7 | 91.8 | 77.4 | 59.4 | 80.8 | 50.0 | 75.9 | 87.0 |
| Adjuvant chemotherapy (all Stage II–III), % | 60.7 | 52.5 | 88.7 | 81.2 | 80.8 | 47.7 | 58.6 | 56.5 |
| Adjuvant chemotherapy (Stage II–III followed at study hospital), % | 75.0 | 55.4 | 95.1 | 89.5 | 100.0 | 86.4 | 77.3 | 65.0 |

Decisions led by the surgeon are reported here rather than adjusted for. \* Computed across all 690 included patients (surgery-first plus neoadjuvant chemotherapy). NA, not evaluated (S8 had fewer than 10 surgery-first Stage II patients). Adjuvant chemotherapy was guided by medical oncologists according to standard guidelines.

Among S6’s Stage II patients who died (n = 9), 1 death occurred within 2 years (median time to death 34.1 months). All 9 patients underwent R0 resection (5 Stage IIA, 4 Stage IIB); 1 had fewer than 16 retrieved lymph nodes (12 nodes) and major postoperative morbidity. Four of the 9 patients were followed at the study hospital, all 4 had recorded adjuvant chemotherapy, and 3 had recurrence documented on imaging. Cause of death, comorbidity, and outside-hospital follow-up records were unavailable in the study database.

### Statistical power and false-positive rate

For a single prespecified surgeon-stage group (unadjusted two-sided α = 0.05, 80% power), detecting a true O/E of 3.0, 2.0, or 1.5 requires about 4, 11, or 39 expected deaths; after two-sided Holm adjustment across 23 tests (α = 0.05/23), these thresholds rise to about 7, 21, and 73 expected deaths. In this cohort, evaluable Stage II cells had 1.5–5.1 expected deaths and Stage III cells had 6.1–16.5. For an average surgeon at this center, detecting a twofold increase in mortality would require about 4 years of accrual in Stage II and 1 year in Stage III without multiple-testing adjustment (about 8 and 2 years after Holm adjustment across 23 tests), whereas detecting a 1.5-fold increase would require about 14 and 4 years unadjusted (27 and 7 years Holm-adjusted).

## Discussion

### Principal findings

In this single-center cohort of 570 surgery-first gastric cancer resections with complete 5-year follow-up, overall 5-year mortality did not differ significantly between the eight surgeons after multiple-testing adjustment, and all Stage III comparisons were compatible with chance. However, stage-stratified auditing benchmarked against all other surgeons identified one large, consistent mortality excess: surgeon S6 in Stage II (9 observed vs 2.27 expected deaths; O/E 3.96, shrunken O/E 2.23), which persisted across every sensitivity analysis.

### Statistical power and volume requirements

The main limitation of surgeon-level survival comparison within a single center is statistical power, not method. The S6 × Stage II excess was detected because it was nearly fourfold (O/E 3.96). Achieving 80% power to detect a still clinically important 1.5-fold increase in mortality depends on event volume rather than calendar time, requiring about 39 expected deaths unadjusted (or 73 after Holm adjustment across 23 tests), equivalent to roughly 240 to 450 Stage II cases at typical 5-year mortality rates. Overcoming this volume constraint in routine practice requires three practical steps: pooling consecutive cases across rolling multi-year windows, aggregating data across regional or national hospital networks, and applying empirical Bayes shrinkage as used in established profiling programs [13,17,18].

### Clinical interpretation and governance

A flagged O/E ratio should serve as a prompt for structured clinical review rather than a direct measure of individual surgical competence. Inadequate lymphadenectomy can shorten survival directly and through stage migration, where a Stage III cancer with too few nodes examined is recorded as Stage II and raises Stage II mortality [26,27], although the S6 × Stage II excess persisted when restricted to patients with at least 16 retrieved nodes. Adjuvant chemotherapy improves survival after D2 gastrectomy in appropriately staged Stage II–III disease [23,28], yet our process audit illustrates a common pitfall of routine-data profiling: S6’s low overall recorded adjuvant chemotherapy rate in surgery-first Stage II–III disease (47.7% vs median 60.7% [range 52.5–88.7%] for the other surgeons) coincided with a low rate of follow-up at the study hospital (50.0% vs median 78.7% [range 59.4–91.8%]); among Stage II–III patients followed at the study hospital, S6’s adjuvant rate was 86.4% (vs median 77.3% [range 55.4–100.0%]). Without external records, we cannot separate incomplete capture of outside-hospital chemotherapy from lower treatment delivery or poorer continuity of care outside the primary center, underscoring the importance of structured institutional follow-up and cross-hospital data linkage. Unmeasured comorbidity, frailty, and performance status remain plausible alternative explanations, as these variables were unavailable in the database and are only partially captured by adjusting flexibly for chronological age. Review of each death, covering cause of death, recurrence pattern and external adjuvant treatment, is the necessary next step, and such signals are best addressed through institutional care pathways: multidisciplinary discussion, referral for perioperative chemotherapy and standards for lymphadenectomy and node examination [29,30].

### Comparison with previous studies

National audits have improved outcomes by feeding risk-adjusted results back to hospitals and surgeons [9,31], and quality improvement directed at variation between providers averts both perioperative and longer-term deaths [32]. In Japan, the National Clinical Database reports risk-adjusted short-term outcomes to institutions without ranking surgeons [33]. Survival has been compared between hospitals with models fitted to those hospitals [10,34], and such comparisons should be read relative to the reference group [35]. The SAH framework extends existing short-term, hospital-level audits by providing a stage-specific, surgeon-level benchmark of 5-year survival that avoids adjusting away surgeon-controlled treatment decisions. Because it relies on pathological TNM stage, routinely recorded demographic and pathology variables, and 5-year vital status, it can be implemented across diverse hospitals and regional registries, and readily extended where comorbidity or ASA grade are also captured. At a minimum, comparing the quality of curative cancer treatment across surgeons, centers, or countries should anchor on these two patient-centered outcomes (stage-specific 5-year survival and major postoperative morbidity), and the same framework can be readily adapted across other disciplines in surgical oncology.

### Limitations

This is a retrospective, exploratory study of eight surgeons in one center and one year (see Prespecification). ASA grade, comorbidity, performance status and clinical stage were not available, and residual confounding, particularly by comorbidity in older patients, may contribute to the S6 result. The primary exact confidence intervals treat predicted risks from the peer-surgeon model as fixed, although bootstrap resampling that incorporated model-estimation uncertainty left the 95% CI for S6 × Stage II above 1 (1.69–8.67). Because 5-year mortality in Stage III was high (43.5%), the Poisson approximations used for the funnel-plot contours and empirical Bayes shrinkage are slightly conservative compared with our exact binary-outcome intervals, contributing to near-zero estimated between-surgeon variation in Stage III (CV 0.01). Overall survival captures all-cause mortality rather than cancer-specific death; because cause of death was unavailable from mortality registry linkage, competing non-cancer deaths could not be separated, particularly among older patients and those with earlier-stage disease. Finally, 5-year survival inevitably reflects surgical care delivered at least 5 years earlier; earlier endpoints, such as 3-year overall or recurrence-free survival, should be evaluated to enable more timely quality improvement feedback.

## Conclusion

A stage-specific comparison of 5-year survival and major postoperative morbidity between surgeons, benchmarked against all other surgeons, can be computed from routinely recorded clinical and pathology data, but a single center can reliably detect only large mortality differences. Within the SAH framework, flagged signals should serve as screening prompts for structured clinical and follow-up review rather than standalone verdicts on surgical competence. Routine implementation requires pooling data across multi-year windows or regional hospital networks with empirical Bayes shrinkage to achieve adequate event volume.

## Data Availability

Individual patient data supporting the conclusions will be made available to qualified investigators upon reasonable request, subject to appropriate data-sharing agreements and ethics approval. Requests for data should be directed to the corresponding author.

## Acknowledgments

We thank the Shanghai Hospital Quality Control Management Affairs Center and the Medical Affairs Department, Medical Quality Control Center, and Clinical Research Center of Ruijin Hospital for institutional and statistical support, and the nursing staff, research coordinators, and surgical team whose clinical work forms the basis of this audit.

## Declarations

### Funding

This research received no specific grant from any funding agency in the public, commercial, or not-for-profit sectors.

### Conflicts of interest

None declared. None of the authors are among surgeons S1 to S8. B.K.S. holds appointments at the Medical Quality Control Center, Ruijin Hospital, and the Shanghai Hospital Quality Control Management Affairs Center, and D.G. at the Shanghai Center for Health Technology Assessment; no institutional position influenced the analysis or its reporting.

### Ethics approval

Approved by the Ruijin Hospital Ethics Committee, Shanghai Jiao Tong University School of Medicine (approval no. 2025-594; 18 September 2025), with individual informed consent waived for this retrospective institutional quality audit.

### Data availability

Statistical methods details are provided in Supplementary File 1, and full Python analysis scripts are available from the corresponding author upon reasonable request. The de-identified patient-level dataset may be made available on reasonable request, subject to institutional approval.

### Author contributions

B.K.S.: conceptualization, development of the SAH framework, methodology, data curation, formal analysis, software, visualization, writing of the original draft. D.G.: study design, data acquisition, data curation, validation, writing of the original draft, review and editing. J.L.: statistical methodology, validation of the survival analyses, project administration including ethics approval, review and editing. C.D., M.Z., R.J., and X.L.: data acquisition; R.J. and X.L. also contributed to study design. E.C.: study design, supervision, resources, critical revision for important intellectual content. All authors approved the final manuscript and agree to be accountable for all aspects of the work.

## Declaration of AI and AI-assisted technologies in the writing process

During the preparation of this work the authors used Claude (Anthropic) and Gemini (Google) in order to assist with language editing, manuscript structuring, formatting of tables and figures, and independent verification of Python analysis code. After using these tools, the authors reviewed and edited the content as needed and take full responsibility for the content of the publication.

## Notes

### Competing Interest Statement

The authors have declared no competing interest.

### Author Declarations

The study was registered at ClinicalTrials.gov (NCT07180966) and was approved by the Ethics Committee of Ruijin Hospital, Shanghai Jiao Tong University School of Medicine (approval no. 2025-594).

### Summary of Updates

This revised version updates the terminology from the SAH Index to the Surgical Assessment in Healthcare framework to reflect separate evaluation of two co-primary outcomes, five-year overall survival and major postoperative morbidity of Clavien-Dindo grade IIIa or higher. It clarifies the cohort of 690 consecutive resections performed by eight high-volume gastrointestinal oncology surgeons, specifies complete five-year vital status ascertainment through national mortality registries, and refines the text, tables, figures, and statistical reporting for journal submission.

